# Pandemic Lock-down, Isolation, and Exit Policies Based on Machine Learning Predictions

**DOI:** 10.1101/2020.04.29.20084707

**Authors:** Theodoros Evgeniou, Mathilde Fekom, Anton Ovchinnikov, Raphael Porcher, Camille Pouchol, Nicolas Vayatis

## Abstract

The widespread lockdowns imposed in many countries at the beginning of the COVID-19 pandemic elevated the importance of research on pandemic management when medical solutions such as vaccines are unavailable. We present a framework that combines a standard epidemiological SEIR (susceptible-exposed-infected-removed) model with an equally standard machine learning classification model for clinical severity risk, defined as an individual’s risk needing intensive care unit (ICU) treatment if infected. Using COVID-19-related data and estimates for France as of spring 2020, we then simulate isolation and exit policies. Our simulations show that policies considering clinical risk predictions could relax isolation restrictions for millions of the lowest-risk population months earlier while consistently abiding by ICU capacity restrictions. Exit policies without risk predictions, meanwhile, would considerably exceed ICU capacity or require the isolation of a substantial portion of population for over a year in order to not overwhelm the medical system. Sensitivity analyses further decompose the impact of various elements of our models on the observed effects.

Our work indicates that predictive modelling based on machine learning and artificial intelligence could bring significant value to managing pandemics. Such a strategy, however, requires governments to develop policies and invest in infrastructure to operationalize personalized isolation and exit policies based on risk predictions at scale. This includes health data policies to train predictive models and apply them to all residents, as well as policies for targeted resource allocation to maintain strict isolation for high-risk individuals.

## 1. Introduction

A key pandemic management strategy involves non-pharmaceutical interventions such as isolation restrictions, or “lockdown” (Koo et al. 2020). In the case of the global COVID-19 pandemic, many countries have adopted these strategies to control the spread of the virus. These interventions can complement pharmaceutical ones (e.g., treatments, vaccines) but also may be the only available tool if pharmaceutical solutions either are not scientifically possible or take significant time to develop. While epidemic models have been used to inform such policies (Flaxman et al. 2020, Ferguson et al. 2020), how to best initiate or relax (and possibly re-initiate) isolation restrictions is unclear. Some approaches rely on immunity tests (Petherick 2020) or on testing and tracing technologies (Wang et al. 2020). We study a different approach that, instead of using “ex post” immunity or diagnostic tests, utilizes “ex ante” predictive technologies such as machine learning, which have been proved successful in other contexts. This approach can be used even in the absence of medical interventions because it relies solely on using data gathered in the pandemic’s early stages to identify factors that affect the severity of symptoms, not on developing and distributing tests, treatments, or vaccines. This approach can complement those relying on immunity and diagnostic tests or be considered independently.

The type of personalized isolation (“confinement”) and exit (“deconfinement”) policy that we study is as follows: First, the clinical risk score (risk of experiencing symptoms severe enough to require an ICU bed) for each individual is predicted^1^. Second, those with predicted scores above a certain threshold are classified as “severe,” or “high risk,” and the remainder are classified as “mild” or “low risk.” Third, those classified into the high-risk group are subject to stricter isolation and protection (“confined”), while those in the low-risk group are placed under softer restrictions or none at all (“released”). In practice, this can be achieved by targeted allocation of resources (e.g., providing masks and other personal protective equipment [PPE], dedicating health support, delivering groceries and other necessities for free, etc.) to the high-risk group, targeted government communication that differentiates between high- and low-risk individuals, and other targeted policies. Policy makers adjust the high vs. low threshold over time to achieve the desired objectives while meeting required constraints. While we chose to minimize the time to complete exit while not exceeding ICU capacity at any point, other formulations of the underlying multi-objective problem are feasible.

The studied policies rely on two key assumptions: First, only a small percentage of the population is in the high-risk category. For example, estimates for France as of spring 2020 showed that the vast majority of the population, or >99% (Salje et al. 2020), will not experience severe symptoms needing an ICU if infected by the SARS-CoV-2 virus. Second, risk prediction models based, for example, on data from early infections can be developed and deployed if the necessary data are available. While the infrastructure required to achieve this during the COVID-19 pandemic appears limited despite the existence of risk prediction models (e.g., Bertsimas (2020) and Urwin et al. (2020)), appropriate policies may enable pandemic management based on the data-driven risk predictions in the future.

The intuition behind the studied policies is as follows: Using COVID-19 data as an example, were one to (i) determine who the ∼1% of severe cases are and (ii) perfectly and temporarily isolate and protect them, the remainder of the population would be able to continue a more or less normal life. In such an ideal scenario, many low-risk people would get infected and would infect others but none would have severe symptoms because those high-risk individuals already have been correctly identified and perfectly protected. The medical system would not be overwhelmed, no one would die (though we note we do not consider long-term health effects for those infected), and both the society and economy would avoid a major shock from the indiscriminate lockdowns implemented in many countries.

In practice, the effectiveness of such a policy would depend on two critical imperfections. First, risk prediction models might occasionally make mistakes, e.g., false positive and false negative errors. Second, isolation would be imperfect as, for example, high-risk individuals who should be isolated may occasionally encounter those infected (e.g., due to PPE shortages, non-compliance, or family situation) and low-risk individuals who would be able to continue normal life may not do so (e.g., due to fear).

We study how these two imperfections impact the effectiveness of the aforementioned personalized pandemic isolation and exit policies. In order to do so, we extended a standard epidemic model, namely a version of the Susceptible-Exposed-Infected-Removed (SEIR) model^2^ (Kucharski et al. 2020), to incorporate personalized predictions of severity risk. Using simulations, we investigated how prediction models for patient severity may inform policy in two scenarios: Amid an ongoing outbreak, as was the case in France when lockdown began March 17, 2020, and when the outbreak has been curbed and progressive loosening of isolation policies (exit, or deconfinement) may take place, as was the case in France beginning May 11, 2020.

Our analysis is based on assuming hypothetical risk prediction models one may be able to develop for a pandemic. These can rely on factors known to affect the severity of symptoms if infected, when such factors exist. For example, existing research indicates differential impact of COVID-19 depending on age, body mass index, hypertension, diabetes and other factors^3^ (Guan et al. 2020), which already have been used in emerging risk models, such as those reported in Bertsimas (2020) and Urwin et al. (2020).

To populate our simulation models, we used available COVID-19 estimates and data from France as of May 2020 (Di Domenico et al. 2020). At the end of lockdown on May 11, there were about 2,750 ICU beds occupied by people with COVID-19, down from a peak of 7,148 against the French health system’s 6,000-bed capacity. We used current estimates with a reproduction number value of ℛ_0_ = 2.9 prior to lockdown, and 1.5 million people who had been immune or infected when it started in France on March 17, 2020 (Salje et al. 2020). We analyzed uncertainty using Approximate Bayesian Computation (Marjoram et al. 2003).

Our simulations led to the following main observations and corresponding implications:

1. **Isolation and exit policies when based on risk-model predictions could be substantially faster and safer**. Utilizing realistic parameter values and a high-quality risk model at the upper end of Bertsimas (2020), simulations indicated that a complete exit from COVID-19 lockdown could be undertaken in three waves over six months, with only 10% of the population being under strict isolation for longer than three months – all without overwhelming the medical system and exceeding ICU capacity. Simulations indicated that without such a model, a complete exit would take 17 months^4^ and 40% of the population would be subject to strict isolation for over a year or ICU capacity would be exceeded four times over. An alternative way to interpret our results is that even with a good risk model, ∼30% of the population still must be strictly isolated for several months. In other words, the “herd immunity” approach some policy makers have endorsed is impossible without (i) a high-quality risk prediction model and (ii) the ability to strictly isolate a substantial portion of the high-risk population. **Implication:** Governments should invest in individual health data infrastructure to make such models implementable at scale in the future. This entails infrastructure that not only collects data on the few thousand people who exhibited symptoms and went to hospitals but also collecting individual medical data on the entire population to obtain health risk predictions for all residents; see Evgeniou et al. (2020) for further discussion on the resultant data policies, privacy, and other related issues. **Disclaimer**: because such data and policies do not exist in most countries, our policy was not suitable for managing COVID-19 in 2020. Rather, we study how personalized policies based on machine learning predictions could improve pandemic management in the future.
2. **Even moderate-quality risk models already could bring measurable improvements**, relaxing isolation for millions of people months sooner while adhering to existing constraints on medical resources. Further, and somewhat surprisingly, even with imperfect models, imperfect but targeted and optimally timed *partial* isolation policies can be *more* efficient than *full non-discriminatory* lockdowns, as such policies allow safer immunity building while better protecting the high-risk population and keeping the average isolation percentage low. **Implication:** For immediate action, focus on a “minimal viable product” data and models that can be used at scale. Even amid the COVID-19 pandemic, data on age, body mass index, and hypertension and diabetes—all of which can be assessed at a nearby pharmacy for all people within weeks—already can be used with a risk model such as in Bertsimas (2020) to inform policies that could be relevant for practice.
3. **Personalized policies based on risk-model predictions are highly sensitive to the protection level of confined people**. Interestingly, the impact of the protection level of deconfined people on simulated outcomes depends on risk model quality. With a high-quality risk model, the optimal policy builds herd immunity^5^, which can be done faster when deconfined people are less, not more, protected. **Implication:** Personalize resource allocation to protect the confined predicted high-risk people: Distribute them masks and other PPE, supply them with food and other necessities for free, prioritize testing those in contact with them, etc. Do not spread resources; practice targeted allocation.
4. Lastly, **whether an individual is classified as high-vs. low-risk changes dynamically over time**: How one is classified depends not only on one’s individual characteristics but also on the state of the epidemic. Our proposed personalized policy combines the epidemic progression with data science principles and optimally adjusts the high-vs. low-risk classification threshold so as to ensure safe and fast confinement and deconfinement over time. **Implication:** A careful communication strategy, adjusted over time, is needed to convey such personalized policies to the public.

We now provide some comments on related work. As we do, Acemoglu et al. (2020) study the benefits of applying differential isolation restrictions within a multi-risk SIR (susceptible-infectious-removed) model. Their risk groups, however, are static—young (20-44), middle (45-64), and old (65+)—so the resultant policies are rather limited. For instance, their optimal fully targeted policy keeps the “old” group (≈ 20% of the population) in isolation essentially indefinitely, waiting for a vaccine to arrive. In contrast, our high vs. low risk assessment depends on the classification threshold in a machine learning model and changes dynamically as the epidemic progresses, allowing for much faster exits. Recall that with a high-quality risk model, the optimal exit takes six months and only 10% of the population is isolated for more than three months.

Our approaches also differ with regard to the multi-objective nature of managing a pandemic. Acemoglu et al. (2020) treat the problem as a weighted objective function and compute an efficient frontier between the economic and health metrics. We treat the problem as a constrained optimization. We optimize one objective (time to remove the isolation restrictions, which represents the socioeconomic goals) subject to a constraint on the availability of ICU beds (representing the health goal).

Gershon et al. (2020) and Duque et al. (2020) employ a similar constrained approach within their settings and objectives, which differ from ours. Similar to Acemoglu et al. (2020), Gershon et al. (2020) utilize static risk groups but they are not solely based just on age and include children, low-risk adults, high-risk adults, and nursing home occupants. The results are similar to Acemoglu et al. (2020) in the sense that the high-risk groups must be isolated indefinitely; under certain conditions, however, the low-risk may not be isolated at all in their model. Our results are similar in that up to 65% of the population should never be isolated. Duque et al. (2020) study the timing of non-targeted shelter-in-place isolation orders. We also study timing but our model isolates, i.e., targets a different (smaller) fraction of the riskiest remaining population at each new epoch.

Targeting based on risk factors is, of course, not the only way: Birge et al. (2020) study spatial targeting and Camelo et al. (2021) study dual targeting based on risk groups and their activities. Such studies are complementary to ours.

We finally mention three companions to the present paper. First, given the large body of literature developed at the beginning of the COVID-19 pandemic, when multiple research articles were produced daily, we also present the detailed review in a “literature appendix” Garin et al. (2021) in addition to the papers discussed above. Second, because our findings are based on numerical simulations, the code is available via GitHub at https://reine.cmla.ens-cachan.fr/boulant/seair, and the algorithmic details for how our model is implemented in the code are provided in Boulant et al. (2020). This forms a “code appendix” to our paper, following the highest standards of reproducible research. Third, to facilitate dissemination of our results for the general public, we created a non-coding demo “simulator” for the differential policies that we study here: https://ipolcore.ipol.im/demo/clientApp/demo.html?id=305. This demo is pre-populated with the parameters for France used in our simulations but one can change the parameters and simulate the pandemic isolation and exit policies for other counties given their respective situations.

## 2. Model

At a high level, our model is a combination of a rather standard SIR-like compartmentalized epidemiological model with an equally standard machine learning binary classification/risk model.

### 2.1. Compartmentalized Epidemiological Model: Extended SEAIR

We start from the standard <u>S</u>usceptible-Infectious-<u>R</u>ecovered, or SIR, model and extend it in two ways. First, we add three additional kinds of compartments:

- <u>E</u>xposed, to account for those who are infected but are neither infectious nor symptomatic;
- <u>A</u>symptomatic, to account for those who have contracted the disease and are infectious but asymptomatic; and
- IC<u>U</u>, for people who experience severe enough symptoms and require intensive care (or, more generally, whatever one may define as severe cases for a pandemic).

Taken together, our model captures a pandemic where a proportion of infectious but initially asymptomatic individuals are assumed to develop mild symptoms and recover naturally, while the rest develop severe symptoms requiring hospitalisation; see Figure 1. Such a separation of exposed individuals based on severity of symptoms has been a recurring aspect of most modelling approaches (whether deterministic or stochastic) by prominent epidemiologists; see, for instance, Di Domenico et al. (2020), Salje et al. (2020), Ferguson et al. (2020), Sonabend et al. (2021). Including the “A” compartment captures the salient feature of a pandemic like COVID-19, where the disease spreads faster as many contagious individuals are unaware of being infectious. Technically, this also facilitates using the parameter estimates from these epidemiological studies.

**Figure 1.**
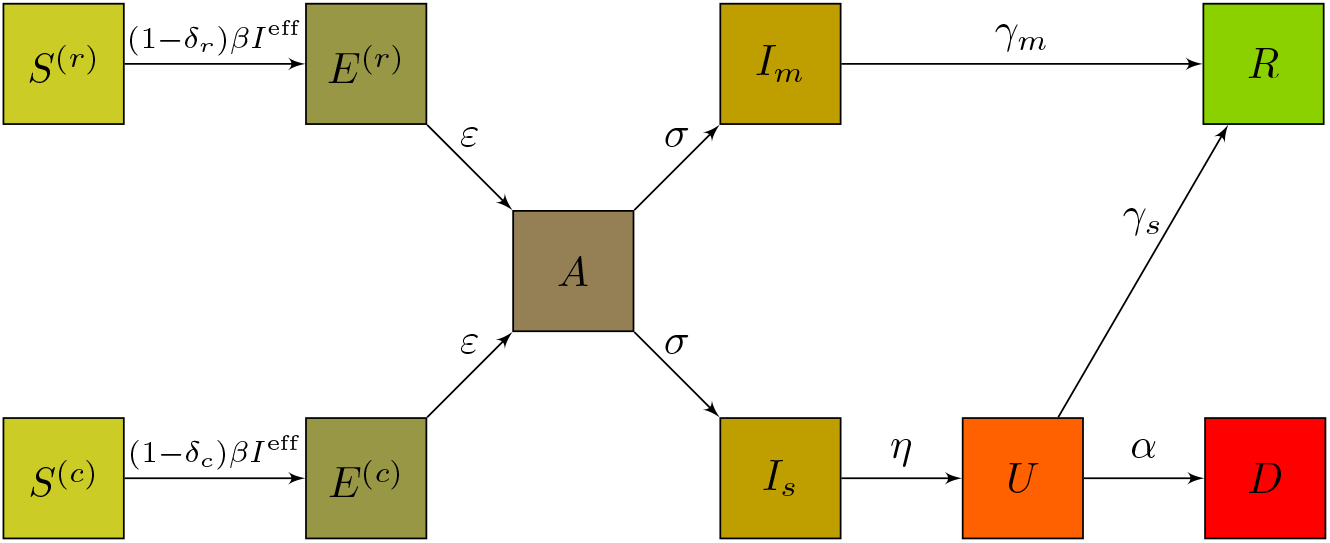
**Simplified schematic of the risk-extended SEAIR model, showing rates of passage from the different compartments: the released**, *S*^(*r*)^, **the confined**, *S*^(*c*)^, **susceptible individuals, the released exposed**, *E*^(*r*)^, **the confined exposed**, *E*^(*c*)^ **the asymptomatic**, *A*, **the infectious with severe symptoms requiring ICU**, *I*_*s*_, **the infectious with milder symptoms**, *I*_*m*_, **the people in ICU beds**, *U*, **as well as those who died from the disease**, *D*, **and those who recovered and are immune**, *R*. **All parameters may be found in Table 1**.

Management studies that explicitly account for asymptomatic cases are rare. For instance, Kaplan (2020) considers the standard SIR model and Acemoglu et al. (2020) consider connected replicas of SIR, while both briefly mention SEIR. Birge et al. (2020), Camelo et al. (2021) start with SEIR and, respectively, split infected individuals into clinical/sub-clinical and confirmed/unconfirmed through testing, the former further divided based on symptoms. These approaches are similar to ours in spirit but suggest that eventually symptomatic individuals may initially be asymptomatic, which necessitates adding the “A”-type compartments.

**Table 1.** Epidemiological parameters used.

| Symbol | Description | Value(s) | Reference |
| --- | --- | --- | --- |
| $\beta$ | transmission rate | computed | |
| $\mathcal{R}_0$ | basic reproduction number | 2.9 | Salje et al. (2020) |
| $\varepsilon$ | waiting rate to viral shedding | 1/3.7 day<sup>-1</sup> | Di Domenico et al. (2020) |
| $\sigma$ | waiting rate to symptom onset | 1/1.5 day<sup>-1</sup> | Di Domenico et al. (2020) |
| $\eta$ | waiting rate from symptom onset to ICU | 1/7 day<sup>-1</sup> | Salje et al. (2020) |
| $\gamma_m$ | recovery rate from mild symptoms | 1/2.3 day<sup>-1</sup> | Di Domenico et al. (2020) |
| $\gamma_s$ | recovery rate for people in ICU | 1/17 day<sup>-1</sup> | Salje et al. (2020) |
| $\alpha$ | mortality rate for people in ICU | 1/11.7 day<sup>-1</sup> | Salje et al. (2020) |

Second, we split each S-E-A-I-R compartment into four categories, with the ICU compartment split in two^6^. Together with a compartment for people who died from the disease, they add up to a total of 23 compartments. The four subcategories correspond to the so-called “confusion matrix” of the machine learning risk prediction model (see Table 2): (i) True Positives, who would experience <u>s</u>evere symptoms upon infection needing and ICU bed, and classified as high-risk and hence <u>c</u>onfined, (ii) False Negatives, who would experience <u>s</u>evere symptoms upon infection needing and ICU bed, but classified low-risk and hence <u>r</u>eleased, (iii) False Positives, who would experience only <u>m</u>ild symptoms upon infection not needing an ICU bed, but classified high-risk and hence <u>c</u>onfined, and (iv) True Negatives, who would experience only <u>m</u>ild symptoms upon infection not needing an ICU bed, and classified low-risk and hence <u>r</u>eleased. How each individual falls into one of these four groups is determined endogenously by our model, as we explain in the next section.

**Table 2.** **Confusion matrix of the risk model**. *f*_*s*_, *f*_*m*_ **denote class-conditional predictive distributions;** *T* **is the classification threshold**, *p* **is the proportion of people with mild symptoms in the population, and** *ρ* **is the proportion of the released population – the decision variable in our model**.

| | Actual, $s$ | Actual, $m$ | |
| --- | --- | --- | --- |
| Model, $s \rightarrow$ confine | True Positives =<br>$(1-p) \times \int_T^1 f_s(\omega) d\omega$ | False Positives =<br>$p \times \int_T^1 f_m(\omega) d\omega$ | $1-\rho$ |
| Model, $m \rightarrow$ release | False Negatives =<br>$(1-p) \times \int_0^T f_s(\omega) d\omega$ | True Negatives =<br>$p \times \int_0^T f_m(\omega) d\omega$ | $\rho$ |
| | $1-p$ | $p$ | |

Because this notation is critically important going forward, we reiterate that the policy we study <u>c</u>onfines those who are predicted to be high risk and <u>r</u>eleases those who are predicted to be low risk. As with any model, our (assumed) prediction model makes mistakes, thus both the confined and released groups contain a mix of actually <u>s</u>evere- and actually <u>m</u>ild-symptom individuals. The sub- and super-scripts *j* = *{s, m}*, for (actually) “severe” vs “mild”, and *i* = *{c, r}*, for “confined” (i.e., predicted severe) vs. “released” (i.e., predicted mild), designate the sub-categories in each compartment. For example, 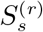 refers to susceptible individuals who are released but will get severe symptoms when infected, *S*^(*r*)^ refers to all released, and so on.

We use *ρ* ∈ [0, 1] as the control parameter of our policy, denoting the proportion of individuals who should be released, i.e., classified in the low-risk group (correctly or incorrectly) and therefore subject to low isolation restrictions. We initially consider a single-release policy, optimizing over a scalar *ρ*, and then extend the analyses to the optimal control problem with multiple releases, optimizing over a vector *{ρ*_0_, *ρ*_1_, …*}* released at times *{*0, *t*_1_, …*}*.

We capture the impact of the differentiated isolation restrictions on people’s behavior using two behavioral parameters: *δ*_*r*_, for the group with low isolation restrictions, i.e., <u>r</u>eleased, and *δ*_*c*_, for the group with high isolation restrictions, i.e., <u>c</u>onfined; 0 ≤ *δ*_*r*_ *< δ*_*c*_ ≤ 1. These parameters capture a level of “protection” and aggregate several factors, such as respiratory and hand hygiene and how much a person has lowered the number of exits from home and social interactions.

Note that how individuals in group *i* = *{r, c}* reduce their chances of contracting the disease depends not only on *δ*_*r*_ and *δ*_*c*_ but also on the proportion of people in each group, *ρ* and 1 − *ρ*. The so-called “contact rates,” *c*_*r*_ and *c*_*c*_ for the released and confined groups, important parameters in “standard” epidemiological models, satisfy 1 − *c*_*i*_ = (1 − *δ*_*i*_) *×* ((1 − *δ*_*r*_)*ρ* + (1 − *δ*_*c*_)(1 − *ρ*)), *i* ∈ *{r, c}*. These parameters have been used in the literature modelling COVID lock-downs, e.g., Djidjou-Demasse et al. (2020), Di Domenico et al. (2020). Our approach is fully aligned with those, as we endogenize contact rates per the preceding equation.

Figure 1 presents the simplified schematic of our SEAIR model, which corresponds to the following set of ordinary differential equations (ODEs), where all parameters are defined in Table 1:

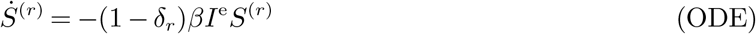

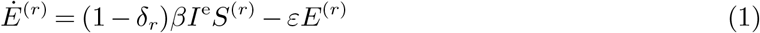

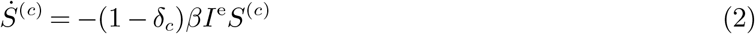

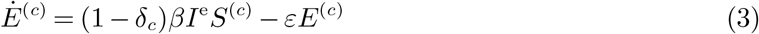

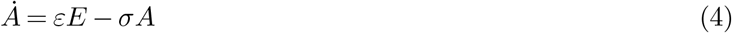

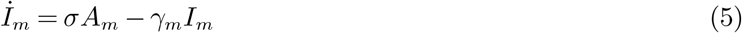

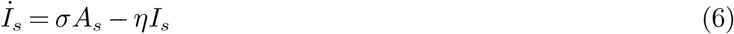

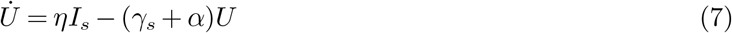

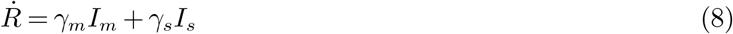

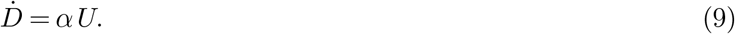

Here, the effective number of contagious people is *I*^e^ = (1 − *δ*_*r*_)(*A*^(*r*)^ + *I*^(*r*)^) + (1 − *δ*_*c*_)(*A*^(*c*)^ + *I*^(*c*)^). As is usual for such models, we consider the so-called *basic reproduction number* ℛ_0_ as measured prior to lockdown. In our model, the situation prior to lockdown corresponds to taking *δ*_*r*_ = *δ*_*c*_ = 0. Then, the transmission rate *β* relates to ℛ_0_ and the other parameters through 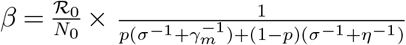. This relation may be derived by linear stability analysis, as in Djidjou-Demasse et al. (2020), using the so-called next generation matrix method (Diekmann et al. 2010).

#### Remark1.

The set of ordinary differential equations (ODEs) and the schematic on Figure 1 present a *simplified* version of our model, reduced from the full set of 23 equations for ease of exposition; e.g., even though the *A* compartment is depicted as a single node on Figure 1 and is presented as a scalar in equation (5), it is actually a four-vector 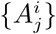, where *i* = *{c, r}, j* = *{s, m}*. The full set of ODEs and the algorithmic details of their solution are presented in the “code appendix” Boulant et al. (2020).

### 2.2. Risk Prediction / Classification Model

Our model necessitates identifying individuals at the highest risk of severity and correspondingly advising them to remain in strict isolation, while relaxing isolation restrictions for lower-risk individuals. Such identification is done in two steps, following the common data science and machine learning approach. For step one, a risk “score” is obtained for each individual, as in Bertsimas (2020), using a logistic regression, random forest, gradient boosting, or the like model. A standard metric to assess the discriminating power of such models is the area under the curve (AUC) of the receiver operating characteristic (ROC) curve (Fawcett 2006). We chose this metric because it is widely used for biomedical applications involving screening populations with some score function (Lasko et al. 2005). The score orders the members of the population from low to high risk and the AUC is often referred to as the rate of concordant pairs (i.e., the fraction of pairs that are correctly compared by the score with respect to their actual risk status). For step two, individuals with risk scores above a certain threshold, *T*, are classified as high risk and are confined, while the rest are classified as low risk and are released. *T* is determined endogenously so the proportion of the released population equals *ρ*, the decision variable in our model.

As mentioned in the introduction, we assume such a risk model would exist in practice. Training such models requires access to non-trivial personalized data and is outside the scope of this paper. Instead, our goal is to evaluate the efficacy of personalized pandemic management given such a model. Therefore, we utilize hypothetical risk models with AUCs that bracket existing COVID-19 models in the literature, e.g., Bertsimas (2020).

To create a hypothetical risk model, let *f*_*s*_ and *f*_*m*_ denote the so-called “predictive distributions” – the PDFs of the risk scores for people with <u>s</u>evere and <u>m</u>ild symptoms, respectively, as predicted by the model. Together with the threshold, *T*, and the proportion of the population with mild symptoms in the population, *p*, these *f*_*s*_, *f*_*m*_ define the model’s confusion matrix, per Table 2. See Clémençon and Vayatis (2007) for other performance measures derived in the context of a control parameter *ρ* applied to a personalized risk model.

As is evident from Table 2, given *ρ, p* and “the model,” i.e., *f*_*s*_, *f*_*m*_, and assuming the risk scores are between 0 and 1, the corresponding classification threshold *T* ^*^ should be selected such that:

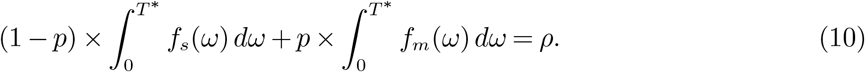

Let *q*^*FP*^ and *q*^*FN*^ denote the model’s false-positive and false-negative error rates, respectively; i.e., 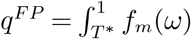 and 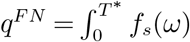. For notational convenience, we omit the dependency of *q*s on *T* ^*^ and, through that, on *ρ*. Then (10) is equivalent to:

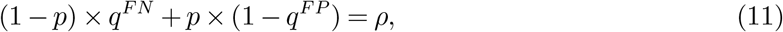

which highlights the key relationship of (any) classification model that we exploit. Selecting a small *ρ* results in a small *T* ^*^, thus a small *q*^*FN*^ as well. Relatively few people will be released but very few of the released would, by mistake, develop severe symptoms. Increasing *ρ* would not only increase *T* ^*^, releasing more people, but it also would increase *q*^*FN*^, exerting a disproportionate impact on people who would require an ICU. An increase in *q*^*FN*^ will be smaller for a higher-quality (higher AUC) model than for a lower-quality one. This is because 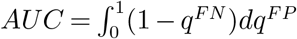, thus a higher AUC implies, ceteris paribus, lower *q*^*FN*^ at a given *T*.

Another important observation from Table 2 is that the released individuals consist of two groups: True Negatives and False Negatives. The latter will experience severe symptoms upon infection, requiring ICU beds. They are not, however, the only group to require ICU beds; because the confinement of the non-released is imperfect (*δ*_*c*_ *<* 1), some of the True Positives will as well. Setting a smaller *ρ*, decreases the number of False Negatives but increases the number of True Positives, leading to a non-trivial relationship between selecting *ρ*, the resultant threshold *T* ^*^, and ICU demand. This relationship depends on the model’s quality (AUC).

The goal is to select *ρ* so that, given the model’s quality, as few people as possible are confined but ICU capacity is not exceeded due to model errors or imperfect confinement.

### 2.3. Connecting Risk and SEIR Models

The risk-model connects with the SEAIR model as follows: For *Q* ∈ *{S, E, A, I, U, R}* and a policy *ρ*, we re-scale the initial risk-independent epidemiologic conditions *Q*_0_ to account for the distribution of people in the four groups:

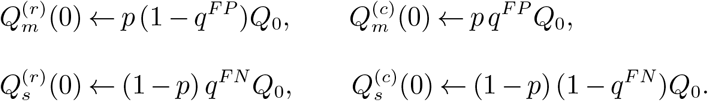

This results in the starting “day 0” conditions for the 23 compartments in the extended SEAIR model, which all depend on *ρ*.

As we investigate policies changing over time, we also update the numbers of people in each compartment when the decision maker increases *ρ* from some value *ρ*_old_ to *ρ*_new_, with corresponding false positive and false negatives rates 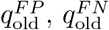 and 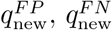, respectively:

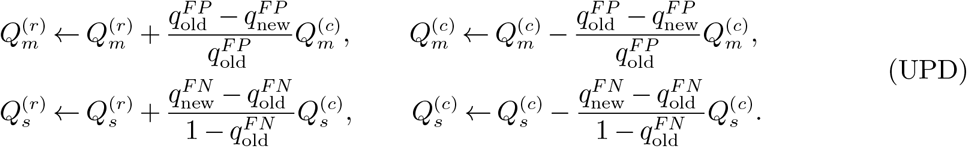

## 3. Estimation of Parameters and Operationalization of Simulations

We simulate the progression of the combined epidemic and risk model discussed in the previous Section in two scenarios: setting “day 0” on March 17, 2020, France’s first day of national lockdown, or on May 11, 2020, the beginning of the lockdown exit. Doing so requires estimating several parameters, listed in Tables 1 and 3. Some of these parameters are inferred from the literature, while others are estimated from data, as we discuss next.

**Table 3.** Simulation parameters used.

| Symbol | Description | Value(s) | Reference |
| --- | --- | --- | --- |
| $N_0$ | total initial number of people in the population | $6.7 \cdot 10^7$ | ∼population of France |
| $S_0$ | total initial number of susceptible people in the population | computed | |
| $E_0$ | total initial number of exposed people in the population | case-dependent | estimated |
| $A_0$ | total initial number of asymptomatic people in the population | case-dependent | estimated |
| $I_0$ | total initial number of infected people in the population | case-dependent | estimated |
| $U_0$ | total initial number of people in ICU | case-dependent | known/estimated |
| $R_0$ | total initial number of immune people in the population | case-dependent | Salje et al. (2020)/estimated |
| $I_{max}$ | hospital capacity for COVID-19 ICU beds | 7 250 | assumed |
| $p$ | proportion with mild symptoms (prior with 90% confidence interval) | 0.9932 [0.9891 – 0.9961] | Salje et al. (2020)/estimated |

### 3.1. Risk model parameters

The class-conditional predictive distributions are modelled as Beta distributions: *f*_*s*_ ∼ Beta(*a*_*s*_, *b*_*s*_) and *f*_*m*_ ∼ Beta(*a*_*m*_, *b*_*m*_). In simulated scenarios, the *no model* refers to *a*_*m*_ = *b*_*m*_ = *a*_*s*_ = *b*_*s*_ = 1. Otherwise, we fix *b*_*s*_ = *a*_*m*_ = 2 and vary^7^ *a*_*s*_ = *b*_*m*_. The *low AUC model* refers to *a*_*s*_ = *b*_*m*_ = 3 (AUC∼75%) and the *high AUC model* refers to *a*_*s*_ = *b*_*m*_ = 5 (AUC∼96%). These parameters are selected so as to bracket the “low” (AUC∼82%) and “high” (AUC∼93%) models from Bertsimas (2020). For sensitivity analyses, we explore the range from *a*_*s*_ = *b*_*m*_ = 2.5 (AUC∼65%) to *a*_*s*_ = *b*_*m*_ = 6.5 (AUC∼99%), further bracketing the range of models that could possibly be available in practice for a disease like COVID-19.

### 3.2. Epidemic model parameters

We estimate the joint distribution of the model parameters by comparing the predictions from our model (at the given parameter values) to the actual data for ICU occupancy obtained from the official portal of the French government: https://dashboard.covid19.data.gouv.fr.

Two general approaches exist for doing so. With the “standard” statistical approach, one splits the data into training and testing sets (sequentially, given the time series nature of the data), learns the model parameters on the training data, and evaluates the predictive accuracy on the testing data. The error structure of the learned model, however, is likely highly non-trivial as the errors are not independent over time. For example, if one SIR-like curve is higher than another early in a time horizon, it must get lower at a later time as fewer susceptible individuals will remain. As a result, this approach could be used to learn the best point estimates but not their joint distribution.

A Bayesian approach can overcome this challenge but it has a noteworth complication: The likelihood function for the resultant prediction errors is also unknown. To deal with this issue, we utilized an Approximate Bayesian Computation (ABC) method (Marjoram et al. 2003), which has been specifically designed for such situations. The ABC method was implemented with the root mean standard error as a distance function (Britton 2010), with a maximum error set at 1,000 ICU beds, which corresponds to the so-called “acceptance rate” of the ABC analyses of about 10%.

Parameters for the initial conditions *S*_0_, *E*_0_, *A*_0_, *I*_0_, *U*_0_ and *R*_0_ depend on the investigated scenario’s “day 0”. The initial number of susceptible individuals, *S*_0_, is computed as *S*_0_ = *N*_0_ − (*E*_0_ + *A*_0_ + *I*_0_ + *R*_0_) − *U*_0_. *N*_0_ and *U*_0_ are known, and an estimate that *E*_0_ + *A*_0_ + *I*_0_ + *R*_0_ ∼ 1.5.10^6^ as of March 17 is available from Salje et al. (2020). How this total splits, however, requires estimation.

To reduce the parameter space, we estimated the total number of exposed, asymptomatic, and infected people, i.e., *E*_0_ + *A*_0_ + *I*_0_, and inferred the number in each state by using the fractions of the mean time spent in each category in the majority population (i.e., people with mild symptoms). More precisely, this corresponds to setting 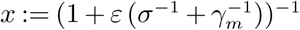 and then:

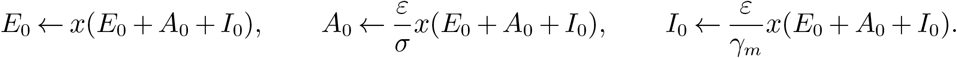

Estimating *E*_0_ + *A*_0_ + *I*_0_ is done jointly with the fraction of individuals with mild symptoms if infected, *p*, and the reduction of contact rates during lockdown, *c*. Note that *c* is not a free parameter in our model (hence it is not listed in Table 3) because, given *δ*_*i*_s and *ρ*, we endogenize *c*_*i*_ for the targeted policies. Because the data to which we fit involve a non-targeted policy, we use a single *c*, corresponding to the single *δ*_*r*_ = *δ*_*c*_ = *δ* with *c* = 1 − (1 − *δ*)^2^.

We used the uniform priors with ranges between 0.5 and 1.4 million for (*E*_0_ + *A*_0_ + *I*_0_) on March 17 and between 65% and 75% for the contact rate during lockdown, *c*, bracketing the estimates from Salje et al. (2020).

We then used the data made available by Salje et al. (2020) to evaluate *p*. This parameter is not given in their work directly but it can be computed as the product of two probabilities: that of being hospitalized upon infection and that of being admitted to the ICU upon hospitalization. Assuming these are independent, the 95% confidence intervals given in Salje et al. (2020) for these two variables translate into a confidence interval to which *p* belongs with a probability of at least 90%, as given in Table 3. This fits a prior Beta distribution with parameters (2265, 15.6).

The number of samples from the prior distributions was set at *n* = 10 000 (resp. 100 000 for robustness when computing means). This led to around 1,000 (resp. 10,000) posterior samples as the acceptance rate was at ∼ 10%. The mean posterior values were found to be *p* ∼ 0.993 and *E*_0_ + *A*_0_ + *I*_0_ ∼ 1200000 (for March 17). The mean posterior value for *c* was found to be *c* ∼ 69.2%, which is consistent with Salje et al. (2020). Reiterating prior discussion, the latter value is unused in our numerical experiments (because we investigate scenarios with differentiated isolation policies) but it is necessary to estimate the joint posterior of *p* and *E*_0_ + *A*_0_ + *I*_0_ from the data about the non-differentiated policy.

#### Remark 2.

Although the obtained mean value of *p* ∼ 0.993 may seem surprisingly high, leaving only ∼ 0.7% probability of dying from a COVID-19 infection, we emphasize that our work does not take into account deaths in nursing homes because those individuals are always confined. Also, in Salje et al. (2020), data show that about 15% of deaths occur during the first day of hospitalization – in other words, those patients are never admitted to the ICU and are not included in an estimate for *p*. That said, we acknowledge that this parameter is critical, hence we also performed sensitivity analyses with respect to *p*; see Appendix A, where we discuss the effect of a significantly lower value *p* = 0.98.

#### Remark 3.

By modelling uncertainty in *p*, we implicitly introduced uncertainty in the risk model as the error rates *q*^*FN*^, *q*^*FP*^ solve equation (11) where *p* is a parameter. We acknowledge that other sources of uncertainty in risk models also could exist^8^.

### 3.3. Simulations with 95% confidence intervals

In all figures showing the evolution of the number of people in the ICU (see Figure 2), the initial condition *E*_0_ + *A*_0_ + *I*_0_ and the proportion of people *p* not requiring ICU admission were sampled according to their posterior distribution. The mean curve of Figure 2 was obtained by taking the average of all the sampled curves, while subsequent 95% confidence intervals were derived by removing the 2.5% and 2.5% upper and lower values for the computed number of ICU beds at each time.

**Figure 2.**
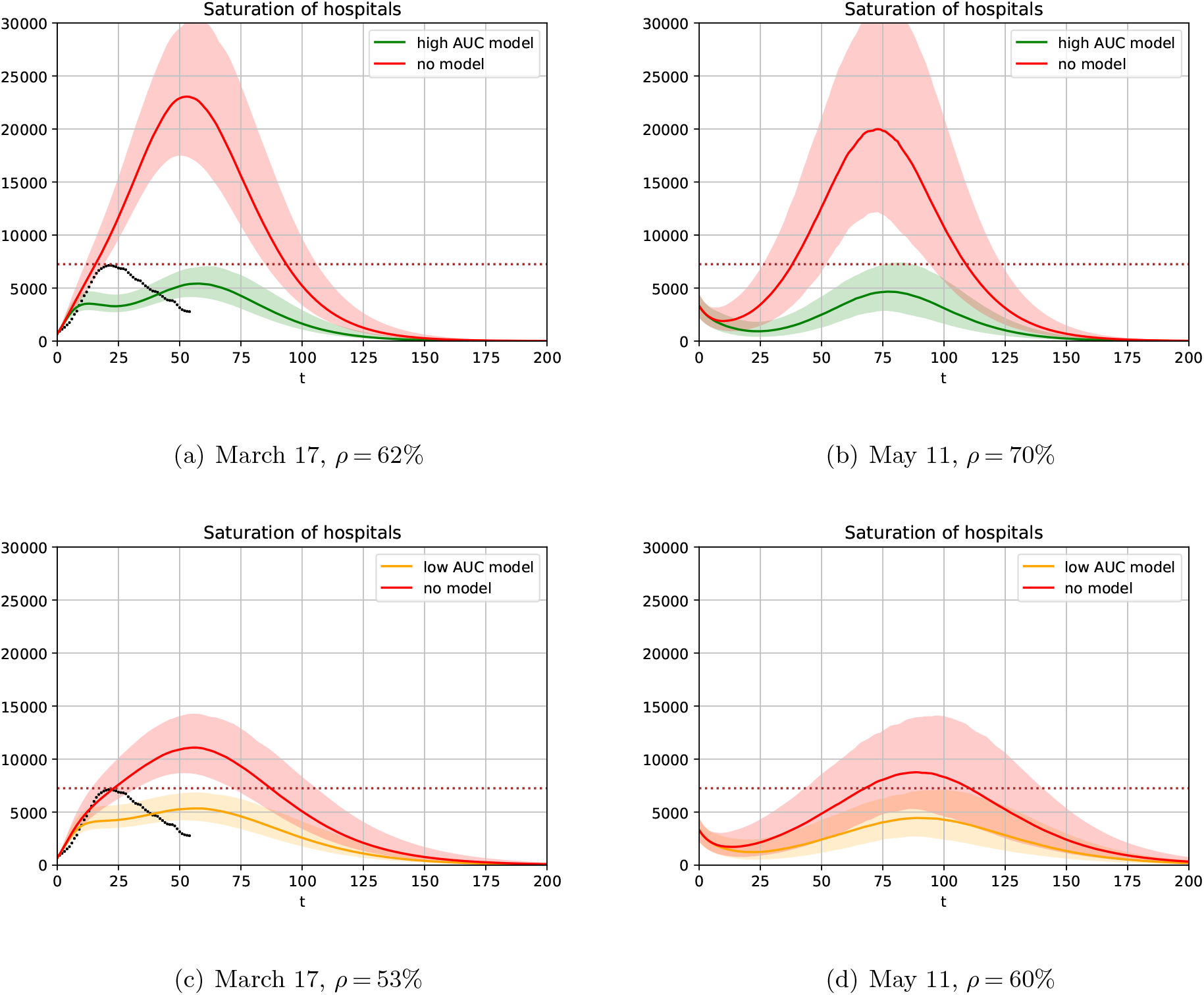
**Number of individuals requiring an ICU bed w.r.t. time** *t* **(days). Left column starts on March 17 (the day of the initial lockdown in France) and right column starts on May 11 (the day when lockdown ended). Dotted line on the left column shows the actual data for France from March 17 to May 11. Top row uses a risk prediction model with high AUC**∼ 95.99**% and bottom row uses a risk prediction model with low AUC**∼ 75.71**%. Without a model**, *ρ* = 45% **for March 17, and** *ρ* = 55% **for May 11**.

### 3.4. Simulations with grid searches

As some numerical experiments (see Figures 3, 7 and Table 4) require grid searches, we did not sample according to the posterior distribution for each scenario. Instead, we computed mean values in order to ease the computational burden. To account for uncertainty, we reduced the number of available ICU beds by the average width of the 95% confidence interval from the corresponding simulations. We then operationalized the simulations as follows:

**Table 4.** **Minimal time (in months) required for all people to exit isolation, starting from March 17 or May 11, depending on** *δ*_*c*_, **model quality and the number of epochs of gradual deconfinement**. *δ*_*r*_ = 0.1

| Four epochs, March 17 |  |  |  | Three epochs, March 17 |  |  |  |
| --- | --- | --- | --- | --- | --- | --- | --- |
|  | High AUC<br>model | Low AUC<br>model | No<br>model |  | High AUC<br>model | Low AUC<br>model | No<br>model |
| $\delta_c = 0.9$ | 6 | >12 | >12 | $\delta_c = 0.9$ | 7 | >12 | >12 |
| $\delta_c = 0.8$ | 8 | >12 | >12 | $\delta_c = 0.8$ | 11 | >12 | >12 |
| $\delta_c = 0.7$ | 10 | >12 | >12 | $\delta_c = 0.7$ | >12 | >12 | >12 |

**Table 4** Minimal time (in months) required for all people to exit isolation, starting from March 17 or May 11, depending on $\delta_c$ , model quality and the number of epochs of gradual deconfinement. $\delta_r = 0.1$
| Four epochs, May 11 |  |  |  | Three epochs, May 11 |  |  |  |
| --- | --- | --- | --- | --- | --- | --- | --- |
|  | High AUC<br>model | Low AUC<br>model | No<br>model |  | High AUC<br>model | Low AUC<br>model | No<br>model |
| $\delta_c = 0.9$ | 6 | 11 | >12 | $\delta_c = 0.9$ | 6 | >12 | >12 |
| $\delta_c = 0.8$ | 7 | 11 | >12 | $\delta_c = 0.8$ | 8 | >12 | >12 |
| $\delta_c = 0.7$ | 9 | 11 | >12 | $\delta_c = 0.7$ | 12 | >12 | >12 |

**Figure 3.**
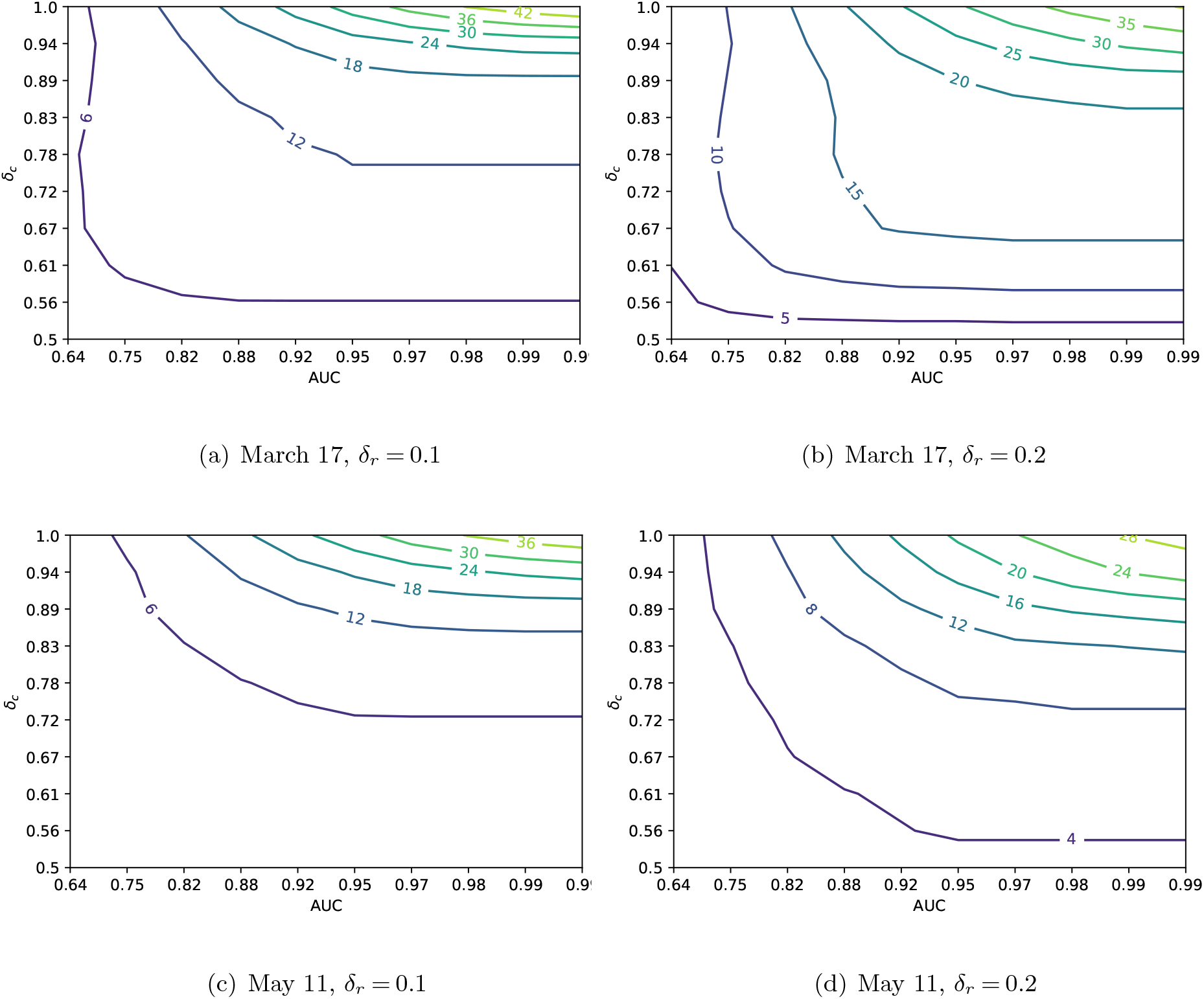
**Difference in maximal percentage of released people without exceeding ICU capacity (7**,**250), compared to the case of not using a risk prediction model, plotted as a function of the AUC of a risk prediction model and the protection level** *δ*_*c*_ **for confined people**. *p* ∼ 0.993 **for all figures. Appendix A presents similar figures with** *p* **decreased to** 0.98, **and ICU capacity increased to 15**,**000**.

- For “day 0” of March 17, 2020: The initial number of utilized ICU beds is known *U*_0_ ∼ 700 and the estimate for the total number *E*_0_ + *A*_0_ + *I*_0_ + *R*_0_ ∼ 1.5.10^6^ is available from Salje et al. (2020). Therefore, we took the average over the posterior and obtained *E*_0_ + *A*_0_ + *I*_0_ ∼ 1200000, resulting in *R*_0_ ∼ 300000 people. Similarly, taking the mean along posterior samples resulted in the estimate of *p* ∼ 0.993. The ODE system was then integrated up until 200 days beyond March 17, 2020.
- For “day 0” of May 11, 2020: Sampling according to the posterior for *p* and *E*_0_ + *A*_0_ + *I*_0_ and integrating the ODE system from March 17 to May 11, we obtained a sample of initial conditions for May 11, of which we took the averages to obtain estimates for all parameters. The ODE system was then integrated up until 200 days past May 11, 2020.

## 4. Results

We present the results of our simulations in four steps. First, we consider a partial exit problem with a single release. Second, we explore the sensitivity of the single release problem. Third, we consider the complete exit problem over multiple release epochs. Finally, we discuss its sensitivity, including the impact of “lockdown fatigue” (Goldstein et al. 2021).

### 4.1. Partial policies with a single release

Figure 2 displays the number of individuals requiring an ICU bed w.r.t. time *t*. The March 17 scenario is in the left column and the May 11 scenario is in the right. Two risk models are considered: a “high” AUC∼ 95.99% (top row) and a low AUC∼ 75.71% (bottom row), bracketing the performance of initial risk models developed for COVID-19 (Bertsimas 2020, Urwin et al. 2020).

In each plot, *ρ* represents the maximal percentage of the population that can be released (i.e., subject to lighter restrictions), which is assumed to correspond to *δ*_*r*_ = 0.1 in such a way that the 95% confidence interval of the number of individuals requiring an ICU bed when using the risk prediction model (green and orange curves) remains below the number of available ICU beds, 7,250. In these first simulations, the rest of the population is confined with more restrictions, *δ*_*c*_ = 0.9. Finally, the red curves show the number of individuals requiring an ICU bed w.r.t. time if the same *ρ* of population is released, but selected at random without any risk prediction model.

#### Initial Lockdown

Figure 2a) shows that on March 17, 2020, a high-AUC model (green curve) allows for *ρ* = 62%. That is, only 38% of the population should be ever confined. In France, which has a population of 67 million, this corresponds to 25 million people. Critically, the remaining 42 million (62%) should never have been confined at all. Figure 2c) shows the same for the low-AUC model (orange curve), which enables *ρ* =53%, or some ∼ 6 million more people in isolation. Perhaps more importantly, without a model, for March 17, *ρ* = 45% – a 17% and 8% difference, respectively, or 5 to 11 million people. In other words, the initial lockdown could have been managed much better if the government had the ability to first train and then utilize a severity risk model at scale.

Figures 2 a) and c) also show actual ICU bed utilization in France by black dots, leading one to wonder why our model requires fewer ICU bed with a partial lockdown than in the actually implemented complete lockdown. This is because the complete lockdown was imperfect: All individuals were able to go shopping, exercise outside, etc., regardless of risk status. From the ABC analyses described in Section 3.2, the estimated reduction in contact rate during the lockdown was ∼ 70%. With a differentiated policy in our model (using the high-AUC model as an example), the 1 − *ρ* = 38% of the population confined with *δ*_*c*_ = 0.9 experiences the contact rate reduction of 1 − 0.1 *×* (0.9 *×* 0.62 + 0.1 *×* 0.38)) = 94%, while the 62% released with *δ*_*r*_ = 0.1 experience the reduction of ∼ 46%. That is, the majority of those who require an ICU bed are better confined and ICU utilization increases more slowly, as depicted.

#### Lockdown exit

Figures 2 b) and d) also present ICU occupancy over time, but for lockdown exit strategies as of May 11. Importantly, unlike what France did on May 11, it is not optimal to release the entire population. Even with a high-AUC model, some 30% should still remain in isolation (40% with the low-AUC model), otherwise ICU capacity would be exceeded in the second wave. Of course, our analyses assume that the released population is subject to *δ*_*r*_ = 0.1 restrictions, while the policy actually implemented in France was aiming for a higher reduction in contact rates. That said, the French government in November 2020 implemented a second lockdown precisely to avoid overwhelming ICU admissions due to the second COVID wave, exactly as we predicted would happen if less than 30%-40% of the population remained in isolation.

These results, however, are for a single release, and a natural question is what one should do with those 30%-40%, which we address in Section 4.3 after discussing the sensitivity analyses.

### 4.2. Sensitivity analyses of the single release policy

Figure 3 displays the results for the March 17 and May 11 scenarios showing the difference between the maximal percentage of people who may be released without exceeding ICU capacity with a risk model, relative to same percentage, but without a risk model. Sensitivity is tested with respect to the discrimination performance of the risk prediction models (AUC) and the degree of isolation of the confined population (*δ*_*c*_). We also alter the degree of isolation for the released population (*δ*_*r*_) across different plots.

As expected, the higher the discrimination of the prediction model, the bigger the difference. However, the degree of isolation has a different impact depending on who is considered: For the confined population, the stricter the isolation (the higher *δ*_*c*_ is), the larger the impact of the risk prediction model. But for the released population (*δ*_*r*_ = 0.1 or 0.2 in Figure 3), the results are more intricate. It is often better to isolate individuals more strictly, except when the risk prediction model is of very high quality and the confined people are in very strict isolation. In those situations, the optimal *ρ* is large enough to achieve “herd immunity” 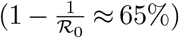, which can be achieved faster if the released population is less protected. The main implication of these analyses is that it is important to both assume in models and encourage in practice stricter isolation practices for the high-risk population – for example, by focusing distribution of PPE and other resources, strictly isolating nursing homes, etc. This not only better protects the high-risk group but also allows for a faster and more efficient exit from the pandemic for the rest.

### 4.3. Complete exit with multiple release epochs, a.k.a. gradual deconfinement

Next, we explore gradual exit strategies, where we optimize both over time and proportion of release with a given fixed number of policy updates *N* while not exceeding a given ICU constraint *M* > 0. In other words, we set a time-optimal control problem, where the variables are the times *t*_1_ ≤ … ≤ *t*_*N*−1_ ≤ *t*_*N*_ and proportions of people released *ρ*_1_ ≤ … ≤ *ρ*_*N*_ with *ρ*_*N*_ = 1, and we aim at minimizing *t*_*N*_, i.e., the moment at which all individuals are released and lockdown is over. The resulting time-optimal control problem is:

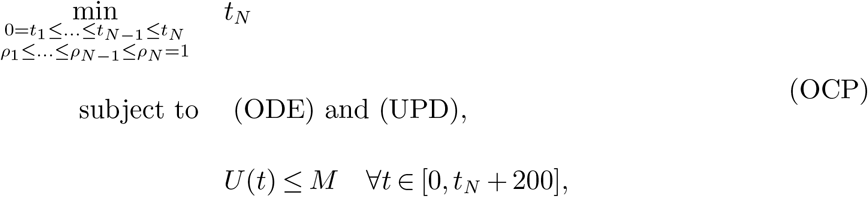

where *U* (*t*) is given by (8) and (ODE) and (UPD) are the systems of equations governing the evolution of the ODE system between the releases and the updating, per Section 2.

To consider practical and realistic scenarios, we solved the resultant optimization dynamic program, allowing releases every 30 days at multiples of 5% of the population, while ensuring the maximum number of utilized ICU beds did not exceed *M* = 5250. We found that implementing the full-blown confidence interval analyses, as in Figure 2, was computationally intractable in the dynamic program setting. However, by observing that the maximum confidence range in Figure 2 was ∼ 2000 beds, we reduced ICU capacity from the “base-case” of 7,250 to *M* = 5,250 to account for uncertainty in ICU demand. For a more realistic initialization, we first solved for the single-release *ρ*^max^, as per Section 4.1 and Figure 2, then fixed *ρ*_1_ = *ρ*^max^. Similarly, for a realistic termination, we ran the ODE model for 200 days after *t*_*N*_ to ensure the ICU constraint is not broken after the entire population is released.

Table 4 shows the minimal number of months, i.e., the optimal solution to problem OCP, to release the entire population for different scenarios (no model, low-AUC model, high-AUC model), while keeping all other parameters constant for three different values of *δ*_*c*_. We considered only gradual releases in three or four epochs; the ICU system was overwhelmed when using only two epochs for most simulations. The main insight is that using no risk model would require more than a year in all scenarios, while an exit with risk-based models would lead to relaxing restrictions for the entire population in as quickly as six months.

Figure 4 shows similar optimal policies for May 11 corresponding to the four epochs from Table 4, and assuming *δ*_*c*_ = 0.9 as in Figure 2. The insights complement those for single release policies: With risk-prediction models, a smaller percentage of the population may need to be confined. Consequently, one also could reach the moment when isolation measures could be lifted sooner.

**Figure 4.**
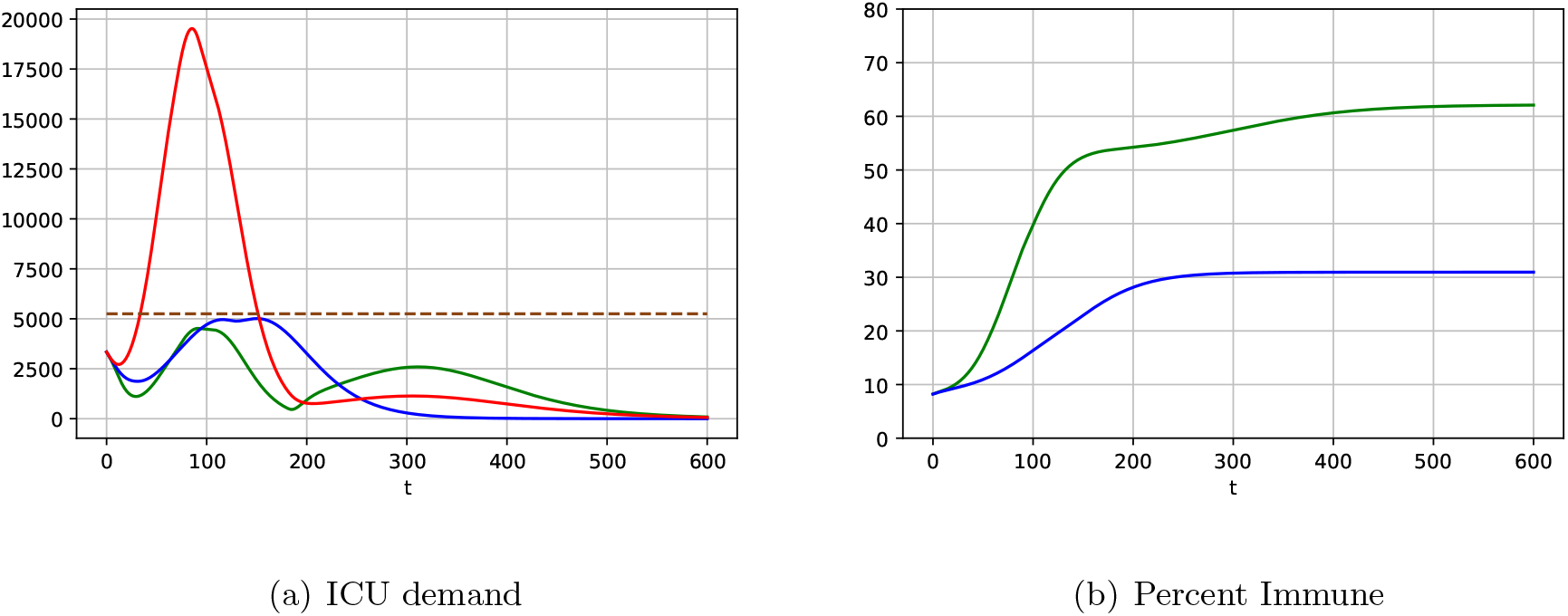
**Examples of gradual schedules of relaxing isolation restrictions with and without model-based risk predictions. High AUC model (green) and no model (red)** *ρ* = [0.65, 0.7, 0.9, 1] **and t** = [60, 90, 180, 600], **no model (blue):** *ρ* = [0.5, 0.55, 0.6, 1] **and t** = [30, 120, 510, 600]. **Vectors t** = [*t*_1_, *t*_2_, …] **give the population release schedules** *ρ* = [*ρ*_1_, *ρ*_2_, …] **as follows:** *ρ*_1_.100% **of the population is released on day** 0, **then** (*ρ*_2_ − *ρ*_1_).100% **are released on day** *t*_1_, **etc**.

For example, using the high-AUC model and without exceeding ICU capacity at any point, 65% of the lowest-risk population could be released on May 11 (“day 0”), followed by another 5% on July 10 (“day 60”), another 20% on August 9 (“day 90”), and the final 10% on November 7 (“day 180”). Resultant ICU demand is shown as a green line on Figure 4a.

Implementing the same exit schedule without a risk model would lead to ICU demand of nearly 20,000 beds (red line). In contrast, a capacity-abiding exit strategy without a model (blue line) would require 17 months to reach full deconfinement. Only 50% of population could be in low isolation on “day 0” (May 11), another 5% on “day 30” (June 10), additional 5% on “day 120” (September 8), and the last 40% only on “day 510” (October 3, 2021), or 11 months later than the similar risk-model-based strategy. Such an extended isolation also would apply to many more people: 10% with the model versus 40% without. For France, this means an additional ∼20 million people in isolation for the additional 11 months.

For both scenarios, Figure 4b shows the percentage of the population that becomes immune over time. Because the model-based policy releases a larger portion of the low-risk population and does so faster, herd immunity is approached, allowing for the ultimate protection against the disease. In contrast, herd immunity is not achieved by a policy without the risk-model: The disease is suppressed but could explode again.

### 4.4. Sensitivity of multiple release policy to lockdown fatigue

The optimal exit policies reported above consist of several exit epochs at which a portion of population is released from isolation, while the rest remain confined. Goldstein et al. (2021) argue that such prolonged restrictions represent a psycho-sociological burden on isolated individuals and could result in “isolation fatigue,” thus diminishing the degree of compliance. Our model can capture this by letting *δ*_*c*_ depend on time, i.e., by defining *δ*_*c*_(**t**) = [*δ*_*c*_(*t*_1_), *δ*_*c*_(*t*_2_), *δ*_*c*_(*t*_3_), *δ*_*c*_(*t*_4_)]. Because the last release epoch is *t*_3_, *δ*_*c*_(*t*_4_) is degenerate, but for technical purposes we set it as *δ*_*c*_(*t*_3_).

We explore two fatigue scenarios: “Low,” where the confined individuals increase their contact rates two-fold by the final release epoch, and “high,” where the increase is three-fold. For simplicity, we assume *δ*_*c*_(*t*) changes linearly over time. That is, the two scenarios correspond to *δ*_*c*_(**t**) = [0.9, 0.85, 0.8, 0.8] and *δ*_*c*_(**t**) = [0.9, 0.8, 0.7, 0.7], respectively.

Figure 5 illustrates two ways in which fatigue impacts our results. First, the black lines show the simulated ICU demand from applying the optimal policy without fatigue (per previous subsection) in a situation where fatigue is present. Clearly, ignoring fatigue could result in substantially exceeding ICU capacity. A policy that ignores fatigue is overly aggressive in relaxing the isolation restrictions as it anticipates the remaining population to be strictly confined. When that is not the case due to fatigue, the policy should be less aggressive early on, which will slow down the build-up of population immunity, also delaying subsequent release epochs.

**Figure 5.**
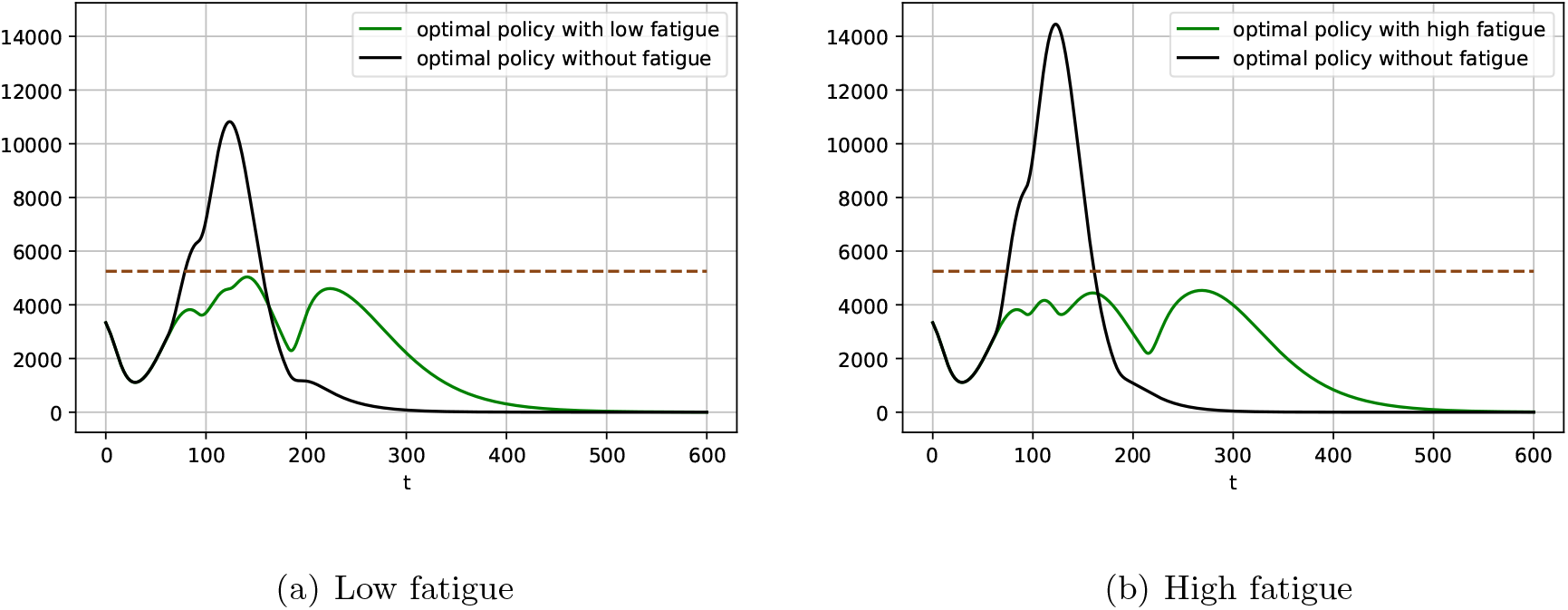
**Examples of gradual schedules of relaxing isolation restrictions with and without model-based risk predictions. High AUC model with fatigue. For the green curves** *ρ* = [0.65, 0.8, 0.85, 1] **and t** = [90, 120, 180, 600] **on the left-hand side figure and** *ρ* = [0.65, 0.7, 0.85, 1] **and t** = [90, 120, 210, 600] **on the right-hand side figure. For both black curves** *ρ* = [0.65, 0.7, 0.9, 1] **and t** = [60, 90, 180, 600]. **Contrary to Figure 4**, *δ*_*c*_ **varies at each release, here** *δ*_*c*_(**t**) = [0.9, 0.85, 0.8, 0.8] **on the left-hand side figure and** *δ*_*c*_(**t**) = [0.9, 0.8, 0.7, 0.7] **on the right-hand side figure**.

Second, the green line shows ICU demand under the optimal policy that takes into account fatigue (and, critically, anticipated fatigue). Contrasting the waves in the two policies illustrates the differences between the optimal policies. For the benchmark, recall that a policy without fatigue releases 65% of population immediately, then releases another 5% on day 60, another 20% on day 90, and the remaining 10% on day 180, i.e., completes the exit in six months.

The optimal policy in the low-fatigue scenario also releases 65% immediately and completes the exit in six months but keeps an additional 10% of people – twice as many – in isolation after day 90, and 15% for days 120-180. The optimal policy in the high-fatigue scenario exacerbates the differences further, keeping an additional 20% of people – three times as many – in isolation beyond day 90, with release completed on day 210 as opposed to 180, and with 15% in isolation for days 120-210.

The driver of the differences is the same in both scenarios. The optimal policy aims at building enough population immunity so that when the highest-risk individuals, many of whom would require ICU treatment upon infection, are released, the infection spreads slowly, allowing the last wave to stretch over an extended period of time, e.g., see Figure 4 (a). Non-compliance due to fatigue restricts the ability to do so, shifting the release further in time, reducing the fraction of release individuals at each epoch, and enlarging the gaps between subsequent epochs.

Generally, the effect of non-compliance with isolation restrictions (due to fatigue or other reasons) is similar to that of lowering the quality of the risk model. Both restrict the ability to target individuals differently given their respective risk factors, thus emphasising the two main messages of our paper: Successful targeted interventions require (i) identifying high-risk individuals and (ii) treating them differently.

## 5. Extensions and Variations

In this section, we discuss several extensions and variations to our model. In particular, because multiple approaches to modelling epidemics have been proposed, it is important to understand how our approach of combining an epidemic and a machine learning model would work in those cases and whether the insights would be qualitatively different.

### 5.1. Nonlinear infection transition term in SIR models

One key modelling phase necessary for the development of any SIR model is to decide at which rate susceptible individuals are infected – that is, to choose a so-called *force of infection λ* in equation 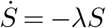. Our model is based on the *density-dependent* approach for transmission, namely *λ* = *βI*, as opposed to the *frequency-dependent* approach for transmission, which reads 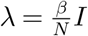, where *N* is the total number of individuals, Begon et al. (2002). The former choice is usually advocated for when the contact rate between individuals increases with density, as is the case for COVID-19.

The situation becomes more intricate when containment comes into play (see Acemoglu et al. (2020) for an in-depth discussion of this scenario). The key point is that in the presence of containment policies, a fine-modelled infection probably should exist between the two approaches. Hence, one way to generalize our model is to interpolate between the two scenarios as Acemoglu et al. (2020) did by introducing an additional parameter *η* ∈ [0, 1] so that 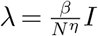.

Importantly, taking *η* = 0, as we do, is a conservative approach because this maximizes the transmission rate among possible *η* choices. In other words, any intermediate value for the parameter *η* will lead to better results in terms of curbing the epidemic, all else equal: faster exits, fewer people in confinement, etc.

### 5.2. Heterogeneity in model parameters – a “contact-matrix”-like approach

Another extension could deal with heterogeneity in model parameters. For instance Baqaee et al. (2020) consider an SIR-like model where each compartment is split by age into five groups: 0-19, 20-44, 45-64, 65-74, and 75+ (Duque et al. (2020) consider a similar split). Then, a susceptible adult of age *a*_1_ who comes into contact with an adult of age *a*_2_ has an instantaneous infection probability of *β* times the probability that the latter adult is infected. The total instantaneous probability of infection is the sum over the expected transmission by contacts of different ages.

Our model has important similarities to and differences from such an approach. We also split each compartment but the split is not static; by choosing *ρ*, we “move” individuals between the groups and the risk model’s ROC curve dictates if such moves are correct or are mistakes. Other than that, the instantaneous infection probability behaves identically; most importantly, it also depends on the behavior and sizes of the released and confined groups, even though we use the same level of *β* at every *ρ*.

A model that combines the contact-matrix approach and ours is not difficult to build. Interestingly, in a context like that of COVID-19, the qualitative outcome of such an exercise is evident. The severity risk increases with age, while the degree of social interactions decreases with age. Therefore, interactions with the elderly will be restricted both because they are confined with *δ*_*c*_ and because they otherwise interact less than young people, i.e., as if *δ*_*c*_ decreases in *ρ*. This, again, makes our results conservative with respect to incorporating the heterogeneous interactions between age groups, and the qualitative results will hold. That said, in contexts other than COVID-19, such combined models may prove valuable.

### 5.3. Alternative epidemic models: stochastic, agent-based, etc

A completely different modelling approach is to use agent-based models that explicitly model each individual whose status evolves according to some probabilistic rules. Such models have been developed to analyze the spread of COVID-19 and other diseases such as seasonal influenza, e.g., Chao et al. (2010, 2020), Koo et al. (2020).

Rahmandad and Sterman (2008) provide an excellent comparison of the ODE- and agent-based models utilizing controlled experiments. Their overarching conclusion is that the differences between the two model types are small. They further note, *“In the realistic situation where policy-makers face time pressure, imperfect data and uncertainty about public behaviour, the wise course may be to use a computationally efficient deterministic compartment model with a broad model boundary, to enable sensitivity and policy tests in time for action, rather than a computationally intensive individual-level model with a constrained boundary and limited ability to carry out parametric and structural sensitivity and policy tests.”* This clearly favors ODE-based models for a new pathogen like SARS-CoV-2.

An additional drawback of agent-based models is their computational load. For instance, Chao et al. (2010) note that a single run of their simulation *“takes about 6 hours (192 hours of total CPU time)”* … *[on a] “cluster of 32 processors.”* This makes a combination of such a model with the optimal control problem like ours hardly feasible. In fact, the other studies that implemented optimal control, such as Acemoglu et al. (2020), Duque et al. (2020), Baqaee et al. (2020), all use SIR-like models. Taken together, these factors suggest that the use of the SIR-like model may be preferred for a study like ours.

### 5.4. Alternative approaches to modelling severity risk

An implicit assumption is that our risk model assigns *individual* risk scores (i.e., a chance that a specific individual, given his or her features, would require an ICU if infected), and that these scores are constant over time. However, this narrow interpretation is not the only possible one, as the following example illustrates: Imagine a household in which two young adults live with their elderly parents. The individual risks for the young adults are low and our model would classify them as released, but an individual risk for an elderly parent may be high, and they would need to be confined. How can our model handle this?

First, because confinement in our model is imperfect, *δ*_*c*_ *<* 1 already implicitly captures multi-generational household situations like the above. Surveys suggest that ∼ 25% of households in the developed world are multi-generational. Combined with the base rate of needing an ICU in single percent, less than ∼ 1% of high-risk individuals would be impacted by this. Because our simulations use *δ*_*c*_ ∈ *{*0.7, 0.9*}*, the multi-generational situations are already captured implicitly.

Second, one can redefine the risk score on a household, rather than individual, level, taking the probability that someone in a household would require an ICU if one of the members gets infected. In practice, this would, of course, require data about the composition of the households and such data would add to the features that the risk classifier uses. Further, with such an approach, one also could implement the dependency of the risk scores on time. Importantly, because the household risk score would be the highest of the individual risk scores, such scores only would decrease over time as high-risk individuals are infected and removed. This, again, makes our results conservative vis-a-vis the efficacy of pandemic management policies with risk predictions.

### 5.5. Alternative objective functions and constraints

Lastly, we comment on the formulation of our optimization model. Unlike some other authors, who evaluate the detailed socio-economic consequences of lockdowns, e.g., Acemoglu et al. (2020), Baqaee et al. (2020), our implicit goal is to design policies that impose lockdowns that affect the smallest number of people and for the shortest period of time. Duque et al. (2020) incorporate this goal explicitly, by minimizing the total number of “people-days” during the lockdown. With our risk model, this is unnecessary because the highest-risk individuals are released last and only when their chance of contracting the disease are small enough because most others have been removed. In other words, it is sub-optimal to confine a lower-risk individual at time *t*_*i*_ if releasing him or her is feasible. This is because releasing strictly increases the number of those removed, allowing for the faster release of the remaining high-risk individuals.

Similarly, we implicitly assume that the release policies are optimized subject to constraints on ICU availability. In practice, however, overall hospital capacity might be reached before that of the ICU. Incorporating this is straightforward: We add two additional compartments before the ICU (see Figure 1), together with the corresponding parameters, which can be easily obtained, e.g., from Salje et al. (2020).

Finally, *δ*_*c*_ in practice may depend on *ρ*. For instance, it may be possible to impose a tougher restriction, 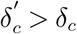, on only a small fraction of the population, but restricting a larger group may only be feasible with 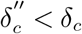. More generally, one could consider *δ*_*c*_(*ρ*) and optimize as we did in Section 4.3 for time-dependent *δ*_*c*_ due to fatigue. This is a plausible extension for our work, but our current results again could be viewed as conservative if one views *δ*_*c*_ as a lower-bound on such a function. Indeed, if the remaining high-risk individuals would be better confined, our model would release more low-risk individuals and sooner.

## 6. Conclusions

Data-driven prediction models, which made large impacts in many areas over the past decades, can help personalized policies for managing epidemic outbreaks. We detailed how prediction models for symptom severity upon infection could be used in epidemic simulations to study the effect of non-pharmaceutical policies, particularly isolation restrictions, during an outbreak. Our core goal is to explore the tension between the prediction model quality and epidemic containment performance. We used COVID-19 data from France as of Spring 2020 as an example and provided sensitivity analyses to understand how different parameters could impact pandemic isolation and exit policies.

Simulations indicated that considering differential relaxation of isolation restrictions depending on predicted severity risk can decrease the immediate percentage of the population in France under stricter isolation by ∼ 5 − 20% relative to not using such risk predictions. Doing so also would speed up a complete exit by several months, directly impacting the lives of millions of people.

We made our simulation engine available to a broad, non-technical audience via an interactive demo that is available at https://ipolcore.ipol.im/demo/clientApp/demo.html?id=305 and is illustrated on Figure 6. The demo is pre-populated with the model parameters for France that we used in this paper but one can input parameters for other countries or regions and experiment with the envisioned policy parameters. We were invited to present our paper to the pandemic management task forces of multiple G7 countries and successfully used this tool to facilitate the dissemination of our results and raise awareness of the promise of personalized data-driven policies for pandemic management.

**Figure 6.**
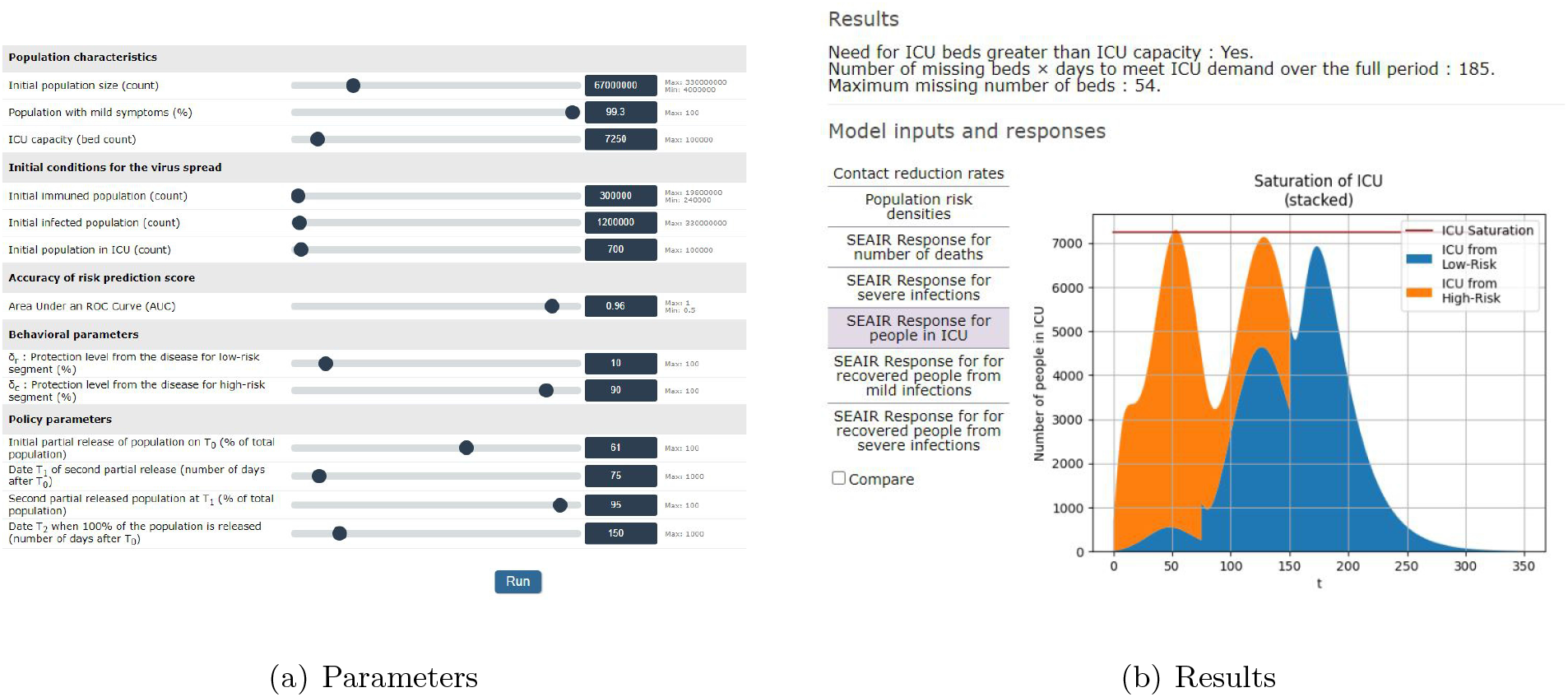
**Screenshots of the demo simulator available here:** https://ipolcore.ipol.im/demo/clientApp/demo.html?id=305.

Sensitivity analyses showed that the qualitative insights from our simulations are robust to changes in risk prediction accuracy, percentage of severe-if-infected cases in the population, availability of resources (e.g., ICUs), and social distancing. Benefits increased when risk prediction accuracy increased, percentage of severe-if-infected cases in the population decreased, availability of resources (e.g., ICUs) increased, and the isolation of high-risk individuals increased.

### 6.1. Limitations and Opportunities for Future Research

Our study is not without limitations, many of which provide opportunities for future research.

First and foremost, all results in this paper use hypothetical risk prediction models based on discrimination ranges in line with early indications from the literature, e.g., Bertsimas (2020), Urwin et al. (2020). To operationalize personalized isolation and exit policies based on risk predictions, governments need to develop policies and invest in infrastructure to enable building and using such models at scale. Critically, this involves both training models on early cases’ data pooled from the epicenter of the pandemic and using models to score entire populations. Currently, the latter presents a bigger problem because even countries with well-organized, single-payer healthcare systems mostly capture data about “sick” individuals (i.e., those who use the healthcare systems) and often lack accurate and recent data for the healthy population. As we discuss in the companion article, Evgeniou et al. (2020), addressing this challenge involves policy matters such as standardisation, privacy, and localisation, as well as technical matters such as transfer and federated learning. All of the above offer opportunities for future research.

Second, epidemic models and their resultant conclusions rely on a number of parameters (e.g., virus incubation and recovery times, basic reproduction number ℛ_0_,) that are uncertain and evolve dynamically, particularly as new virus variants emerge Alban et al. (2020). The resultant policies are therefore contingent: Observing an ICU demand that is closer to an upper boundary of the confidence interval may require the next wave to be delayed or involve a smaller release percentage than our current simulations, built from day zero, suggest. Building models that specifically account for a potential shift in these parameters is an interesting future research direction.

Third, practical policy decisions for a given pandemic require careful context-specific robustness analysis of, for example, the benefit of combining isolation restrictions with other policies such as test-based ones, e.g., Petherick (2020), Wang et al. (2020), or vaccination based ones, e.g., Sonabend et al. (2021). Designing such combined policies is another fruitful research direction.

Finally, risk-predictions based policies using epidemic simulations should be developed taking into account behavioral aspects that may help or hurt model predictions and policy actions: ethical issues, fear, widespread non-compliance to isolation measures, and the like. Behavioral operations studies could inform model building and guide lockdown policy implementation in isolation or in conjunction with other mechanisms. For example, governments in many countries have encouraged vaccination by implementing a de-facto differential isolation policy for non-vaccinated individuals.

In conclusion, our simulations show that combining prediction models using data science and machine learning principles with epidemiological models may improve outbreak management policies. Governments, therefore, should make appropriate investments in the data infrastructure necessary to implement such models at scale and consider their predictions when developing pandemic isolation (confinement) and exit (deconfinement) policies.

## Data Availability

No data. Code is available at https://reine.cmla.ens-cachan.fr/boulant/seair

## Acknowledgments

The authors are grateful to Rams`es Djidjou-Demasse for detailed exchanges about the model of his team Djidjou-Demasse et al. (2020) and to Amaury Lambert and Pierre-Yves Massé for the interesting discussions. We also thank Olivier Boulant for his help in making the code available on GitHub. ADD LINK HERE

## Appendix A: Further sensitivity analyses for a single release

We here gather additional sensitivity analyses in the case of a single release, presented in Figure 7, with respect to key parameters of the model: the percentage *p* of the population with severe symptoms upon infection plays a crucial role, which we decrease from the posterior average ∼ 0.993 to 0.98, and the ICU capacity, which we increase from 7250 to 15000. Other parameters are same as in Figure 3.

**Figure 7.**
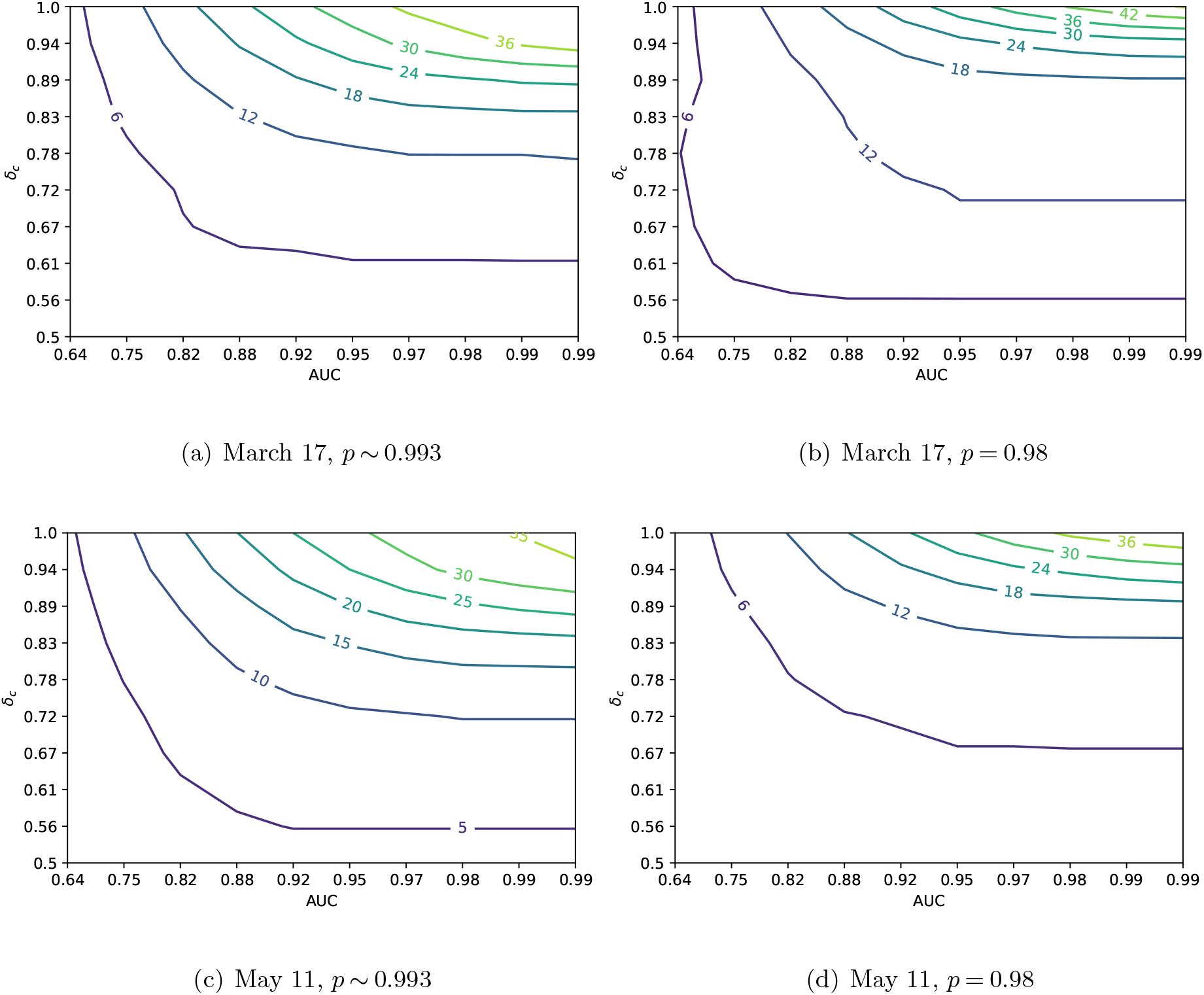
**Difference in maximal possible percentage of released people without exceeding the ICU capacity, compared to the case of not using a risk prediction model, plotted as a function of the AUC of a risk prediction model and the protection level** *δ*_*c*_ **for confined peoplections**. *δ*_*r*_ = 0.1 **for all figures**.

First, as expected, the lower *p*, the less the impact of a risk prediction model keeping the risk-model AUC constant. Given the limited ICU – and possibly other – resources, a smaller *p* allows for a smaller range of percentages of the population being released, making all differences between policies smaller in absolute terms. Second, when we compare Figures 3 (a) and (c) with Figures 7 (a) and (c), where the only difference is in the total ICU capacity, we see that the more ICUs available the larger the impact of using personalized policies, keeping everything else constant. More available resources allow for a larger range of percentage of released people making the differences between policies – risk based vs. not – larger in absolute terms. Note that in all cases a risk prediction model approach allows for confining fewer people; this is consistent with the value of information-related arguments, as any test provides information that can be beneficial assuming everything else (including behavioral aspects) kept constant.

## Footnotes

1 The individual prediction is not critical for our analyses; an alternative prediction target could, for example, be the risk that someone in a household develops severe symptoms. See Section 5.

2 See Section 5 for discussion of alternative modeling frameworks and their pros and cons for our problem.

3 Given that the dependency of the COVID-19 clinical outcomes on age attracted enormous media attention, we note that policies based only on age may be unnecessarily simplistic. For example, ∼ 10% (source: statista.com) of the French population is older than 75, yet only ∼ 1% of the entire population will require an ICU bed if infected. It is thus not practical to implement differential isolation and exit policies based just on age. Instead one may need to only consider models that capture more nuanced and accurate patterns using multiple features, not just age.

4 Such long and massive lockdowns in the absence of a reasonable risk model are consistent with the literature, e.g., Acemoglu et al. (2020) propose isolating all seniors for 18 months, and even that hinges on the assumption that a vaccine will be developed by then. Our analyses do not rely on the availability of vaccines.

5 DISCLAIMER: We do not advocate for herd immunity but we emphasize that building herd immunity may be optimal in some cases, notably when high-risk individuals are well-protected and the model is of a sufficient quality to identify such individuals with a high-enough accuracy. Without these, a herd immunity strategy that does not overwhelm the medical system is impossible.

6 By definition, only severe cases go to the ICU but they can come from two categories: those classified by the model as high risk (True Positives) and those classified as low risk (False Negatives).

7 The symmetric ROC models we considered are not the only possible approach. While working on this paper, we explored multiple non-symmetric settings and observed no qualitative impact on the simulation results.

8 Uncertainty in risk models (i.e., in risk predictions and even in the classification threshold) could be due sampling bias (e.g., clinical trial data do not reflect population data, electronic medical records fail to account for the part of the population never admitted to the hospital) or methodological bias (e.g., model misspecification, suboptimal machine learning/statistical method). To provide statistical guarantees on the risk estimators, it may be necessary to compute confidence bands on the estimated ROC curve, which will then lead to explicit confidence bands on the risk model. Typical approaches to derive confidence bands are to perform error propagation on distribution parameters (in a parametric framework) or to generate several ROC curves through resampling and provide some bootstrap estimate of the confidence band (Macskassy et al. 2005). Resampling strategies may include label flipping (prediction uncertainty), sample perturbation, or shifting (sampling bias). Lastly, the quality of a prediction model was assessed by AUC; other measures also exist, Reid and Williamson (2011).

## References

Acemoglu D, Chernozhukov V, Werning I, Whinston MD (2020) A multi-risk sir model with optimally targeted lockdown (27102), URL http://dx.doi.org/10.3386/w27102.

Alban A, Chick S, Dongelmans D, Sluijs A, Wiersinga W, Vlaar A, Sent D (2020) Icu capacity management during the covid-19 pandemic using a stochastic process simulation. SSRN Electronic Journal URL http://dx.doi.org/10.2139/ssrn.3570406.

Baqaee D, Farhi E, Mina M, Stock JH (2020) Policies for a second wave. Brookings Papers on Economic Activity.

Begon M, Bennett M, Bowers RG, French NP, Hazel S, Turner J (2002) A clarification of transmission terms in host-microparasite models: numbers, densities and areas. Epidemiology & Infection 129(1):147–153.

Bertsimas D (2020) Mortality risk calculatory. URL https://www.covidanalytics.io/calculator.

Birge JR, Candogan O, Feng Y (2020) Controlling epidemic spread: Reducing economic losses with targeted closures. University of Chicago, Becker Friedman Institute for Economics Working Paper (2020-57).

Boulant O, Fekom M, Pouchol C, Evgeniou T, Ovchinnikov A, Porcher R, Vayatis N (2020) SEAIR Framework Accounting for a Personalized Risk Prediction Score: Application to the Covid-19 Epidemic. URL https://ipolcore.ipol.im/demo/clientApp/demo.html?id=305.

Britton T (2010) Stochastic epidemic models: a survey. Mathematical biosciences 225(1):24–35.

Camelo S, Ciocan DF, Iancu DA, Warnes XS, Zoumpoulis SI (2021) Quantifying the benefits of targeting for pandemic response. medRxiv URL http://dx.doi.org/10.1101/2021.03.23.21254155.

Chao DL, Halloran ME, Obenchain VJ, Longini Jr IM (2010) Flute, a publicly available stochastic influenza epidemic simulation model. PLoS Comput Biol 6(1).

Chao DL, Oron AP, Srikrishna D, Famulare M (2020) Modeling layered non-pharmaceutical interventions against sars-cov-2 in the united states with corvid. Working paper 6(1).

Clémençon S, Vayatis N (2007) Ranking the best instances. Journal of Machine Learning Research 8(88):2671–2699, URL http://jmlr.org/papers/v8/clemencon07a.html.

Di Domenico L, Pullano G, Sabbatini CE, Boëlle PY, Colizza V (2020) Expected impact of lockdown in île-de-france and possible exit strategies URL http://dx.doi.org/10.1101/2020.04.13.20063933.

Diekmann O, Heesterbeek J, Roberts MG (2010) The construction of next-generation matrices for compartmental epidemic models. Journal of the Royal Society Interface 7(47):873–885.

Djidjou-Demasse R, Michalakis Y, Choisy M, Sofonea MT, Alizon S (2020) Optimal covid-19 epidemic control until vaccine deployment URL http://dx.doi.org/10.1101/2020.04.02.20049189.

Duque D, Morton DP, Singh B, Du Z, Pasco R, Meyers LA (2020) Timing social distancing to avert unmanageable covid-19 hospital surges. PNAS 117(33):19873 – 19878.

Evgeniou T, Hardoon R D, Ovchinnikov A (2020) Leveraging ai to battle this pandemic — and the next one. Harvard Business Review.

Fawcett T (2006) An introduction to roc analysis. Pattern Recognition Letters 27(8):861–874, ISSN 0167-8655, URL https://doi.org/10.1016/j.patrec.2005.10.010, rOC Analysis in Pattern Recognition.

Ferguson N, Laydon D, Nedjati Gilani G, Imai N, Ainslie K, Baguelin M, Bhatia S, Boonyasiri A, Cucunuba Perez Z, Cuomo-Dannenburg G, et al. (2020) Report 9: Impact of non-pharmaceutical interventions (npis) to reduce covid19 mortality and healthcare demand.

Flaxman S, Mishra S, Gandy A, Unwin H, Coupland H, Mellan T, Zhu H, Berah T, Eaton J, Perez Guzman P, et al. (2020) Report 13: Estimating the number of infections and the impact of non-pharmaceutical interventions on covid-19 in 11 european countries.

Garin M, Limnios M, Nicolaï A, Bargiotas I, Boulant O, Chick S, Dib A, Evgeniou T, Fekom M, Kalogeratos A, Labourdette C, Ovchinnikov A, Porcher R, Pouchol C, Vayatis N (2021) Epidemic models for covid-19 during the first wave from february to may 2020: a methodological review.

Gershon D, Lipton A, Levine H (2020) Managing covid-19 pandemic without destructing the economy.

Goldstein P, Levy Yeyati E, Sartorio L (2021) Lockdown fatigue: The diminishing effects of quarantines on the spread of covid-19. URL https://doi.org/10.21203/rs.3.rs-621368/v1.

Guan W, Ni Z, Hu Y, Liang W, Ou C, He J, LL (2020) Clinical characteristics of coronavirus disease 2019 in china. New England Journal of Medicine URL http://dx.doi.org/10.1056/NEJMoa2002032.

Kaplan EH (2020) Covid-19 scratch models to support local decisions. Forthcoming, Manufacturing Services Operations Management. URL http://dx.doi.org/10.2139/ssrn.3577867.

Koo JR, Cook AR, Park M, Sun Y, Sun H, Lim JT, Tam C, Dickens BL (2020) Interventions to mitigate early spread of sars-cov-2 in singapore: a modelling study. The Lancet Infectious Diseases ISSN 1473-3099, URL https://doi.org/10.1016/S1473-3099(20)30162-6.

Kucharski AJ, Russell TW, Diamond C, Liu Y, Edmunds J, Funk S, Eggo RM, Sun F, Jit M, Munday JD, Davies N, Gimma A, [van Zandvoort] K, Gibbs H, Hellewell J, Jarvis CI, Clifford S, Quilty BJ, Bosse NI, Abbott S, Klepac P, Flasche S (2020) Early dynamics of transmission and control of covid-19: a mathematical modelling study. The Lancet Infectious Diseases ISSN 1473-3099, URL https://doi.org/10.1016/S1473-3099(20)30144-4.

Lasko TA, Bhagwat JG, Zou KH, Ohno-Machado L (2005) The use of receiver operating characteristic curves in biomedical informatics. Journal of Biomedical Informatics 38(5):404–415, ISSN 1532-0464, URL https://doi.org/10.1016/j.jbi.2005.02.008, clinical Machine Learning.

Macskassy SA, Provost F, Rosset S (2005) Roc confidence bands: An empirical evaluation (New York, NY, USA: Association for Computing Machinery), ISBN 1595931805.

Marjoram P, Molitor J, Plagnol V, Tavaré S (2003) Markov chain monte carlo without likelihoods. Proceedings of the National Academy of Sciences 100(26):15324–15328.

Petherick A (2020) Developing antibody tests for sars-cov-2. The Lancet, World Report 395, ISSN 1101-1102, URL https://doi.org/10.1016/S0140-6736(20)30788-1.

Rahmandad H, Sterman J (2008) Heterogeneity and network structure in the dynamics of diffusion: Comparing agent-based and differential equation models. Management Science 54(5):iv–1036.

Reid MD, Williamson RC (2011) Information, divergence and risk for binary experiments. Journal of Machine Learning Research 12(22):731–817.

Salje H, Tran Kiem C, Lefrancq N, Courtejoie N, Bosetti P, Paireau J, Andronico A, Hozé N, Richet J, Dubost CL, Le Strat Y, Lessler J, Levy-Bruhl D, Fontanet A, Opatowski L, Boelle PY, Cauchemez S (2020) Estimating the burden of sars-cov-2 in france ISSN 0036-8075, URL http://dx.doi.org/10.1126/science.abc3517.

Sonabend R, Whittles LK, Imai N, Perez-Guzman PN, Knock ES, Rawson T, Gaythorpe KA, Djaafara BA, Hinsley W, FitzJohn RG, et al. (2021) Non-pharmaceutical interventions, vaccination, and the sars-cov-2 delta variant in england: a mathematical modelling study. The Lancet.

Urwin SG, Kandola G, Graziadio S (2020) What prognostic clinical risk prediction scores for covid-19 are currently available for use in the community setting?

Wang CJ, Ng CY, Brook RH (2020) Response to COVID-19 in Taiwan: Big Data Analytics, New Technology, and Proactive Testing. JAMA 323(14):1341–1342, ISSN 0098-7484, URL http://dx.doi.org/10.1001/jama.2020.3151.

